# How To Enhance Empathy Nursing Students In Education: Literature Review

**DOI:** 10.1101/2022.01.01.22268600

**Authors:** Bhakti Permana, Moses Glorino Rumambo Pandin

## Abstract

**Introduction:** The tendency of reducing student empathy requires efforts to improve it through education or training. The purpose of this review is to identify education, learning, or teaching that is used to increase student empathy.

**Method:** The design used a Literature Systematic Review. Articles were conducted in three electronic databases guided by The PRISMA. Articles published in English and in 2019 to 2021.

**Results:** The result obtained 20 articles and will be reviewed. Eighteen articles show an effect or relationship between intervention and empathy. Training materials that can increase student empathy are the concept of empathy, communication, mindfulness, and transcultural nursing education. Education and training using patients/patient simulations to help improve student empathy, namely: expert patients, ECARE Program, polypharmacy effects; clinical Simulation of Inpatients, Lectures in class, Virtual Dementia Tour, living in poverty, and games for team interaction. Another method is through the KSS module, mannequin simulator experience, and peers.

**Conclusion:** Education and training on the concepts of empathy, communication, meditation, and cultural competence using Simulation and immersion methods with patients, being like patients, using mannequins, or interacting with vulnerable groups can increase student empathy. Faculty and lecturers can apply experiential learning methods with Simulation and immersion in learning or training courses.

## 1. Introduction

Nursing students are often involved in nursing services for patients when conducting the clinical nursing practice in hospital. Students may not be ready to care and have negative behaviors from patients like being afraid, incapable, helpless, anxious, sad, low self-esteem, unable to manage their emotions, and maintaining patient space (Kapucu & Bulut, 2018; Lin et al., 2017). Feeling under pressure, failing to use therapeutic communication skills, a lack of courage, eastern cultural influences, and emotional management experience are all aspects that affect nursing students’ views (Lin et al., 2017). Empathy problems, together with negative attitudes and stigma, are an issue for health care improvement. (McKenna et al., 2012).

Empathy is the process of placing oneself in another person’s shoes, viewing events from a different perspective, understanding and feeling that others’ emotions and ideas, and explaining this condition to him (Cunico et al., 2012; Paola Ferri et al., 2017). The dimensions of empathy are cognitive, emotional, and behavioral. The emotional dimension shows potential for empathy. The cognitive and behavioural dimensions are the ability to empathize.

Empathy in health services is an essential component of a quality therapeutic and nursing relationship (Hojat, 2016). Empathy for a health professional will reduce depression, anxiety, distress; and increase emotional well-being, satisfaction, and adherence to treatment regimens (Hojat & Hojat, 2016) and can be used to carry out patient-centered care and interactions (Omid et al., 2018). Therefore, high empathy is essential for the professional development of students in preparing for the medical profession (Sapiro et al., 2004).

Several studies have shown that students’ empathy levels can decrease during nursing education (Ward et al., 2012), the average medical profession student is lower than non-medical students, and 18.2% of all medical program students (n = 412) showed the lowest level of empathy (Sobczak et al., 2021). It requires programs to implement educational interventions to increase students’ empathy levels.

The importance of empathic skills and the tendency to decrease empathy for students requires a nursing education process to develop empathy skills. Training in communication skills, experiential learning, and medical interprofessional education interventions, and simulation-based education, are all effective techniques for increasing students’ empathy levels (Batt-Rawden et al., 2013; Bearman et al., 2015). For this reason, it is necessary to explore the latest developments in empathy education interventions to increase nursing students’ empathy.

## 2. Review Methods

### 2.1 Study Objectives

The goal of this review is to identify the education, learning, or teaching that is used to increase the empathy of nursing students.

### 2.2 Eligibility Criteria

The type of research used is experimental design, quasi-experimental and mixed methods with quasi-experiments published in English between January 2019 to 2021. The type of the participant is nursing students regardless of gender, age, or country. The type of intervention chosen is an educational intervention designed to increase the level of empathy of nursing students. The type of outcome measure is quantitative data that is objectively measured or self-reported about empathy.

### 2.3 Search Strategy

Search articles or journals using keywords. The keywords used are “Nursing student” OR “Nurse student” AND Education OR Training OR Learning AND Empathy. The literature search used three databases, namely Scopus, Scient Direct, and Sage journals. In addition to searching for articles using keywords, the characteristics of the journal are also determined, namely publications in 2019 and above, full text, and English.

### 2.4 Study Selection

The identified articles are entered into the Mendeley database, and then duplicate titles are identified and deleted. Potential articles will be screened by title and abstract based on inclusion criteria.

### 2.5 Critical Appraisal

Articles that have met the criteria will then be assessed for quality using the standard assessment format from the JBI Critical Appraisal checklist and MMAT. The selection of the assessment format is based on the design of each quasi-experiment article using the JBI Critical Appraisal Checklist for Quasi-Experimental Studies and Mixmethod using the Mixed Methods Appraisal Tool (MMAT). Critical appraisal assesses eligible studies if the research score is at least 60% of the cut-off point value. Screening results showed that 20 articles achieved a score higher than 60%.

### 2.6 Data Collection Process

Articles that meet the criteria and study quality will be tabulated using a table of researchers and year of publication, objectives, design, sample, variables, instruments, analysis, intervention, and results (Table 1).

**Tabel 1.**
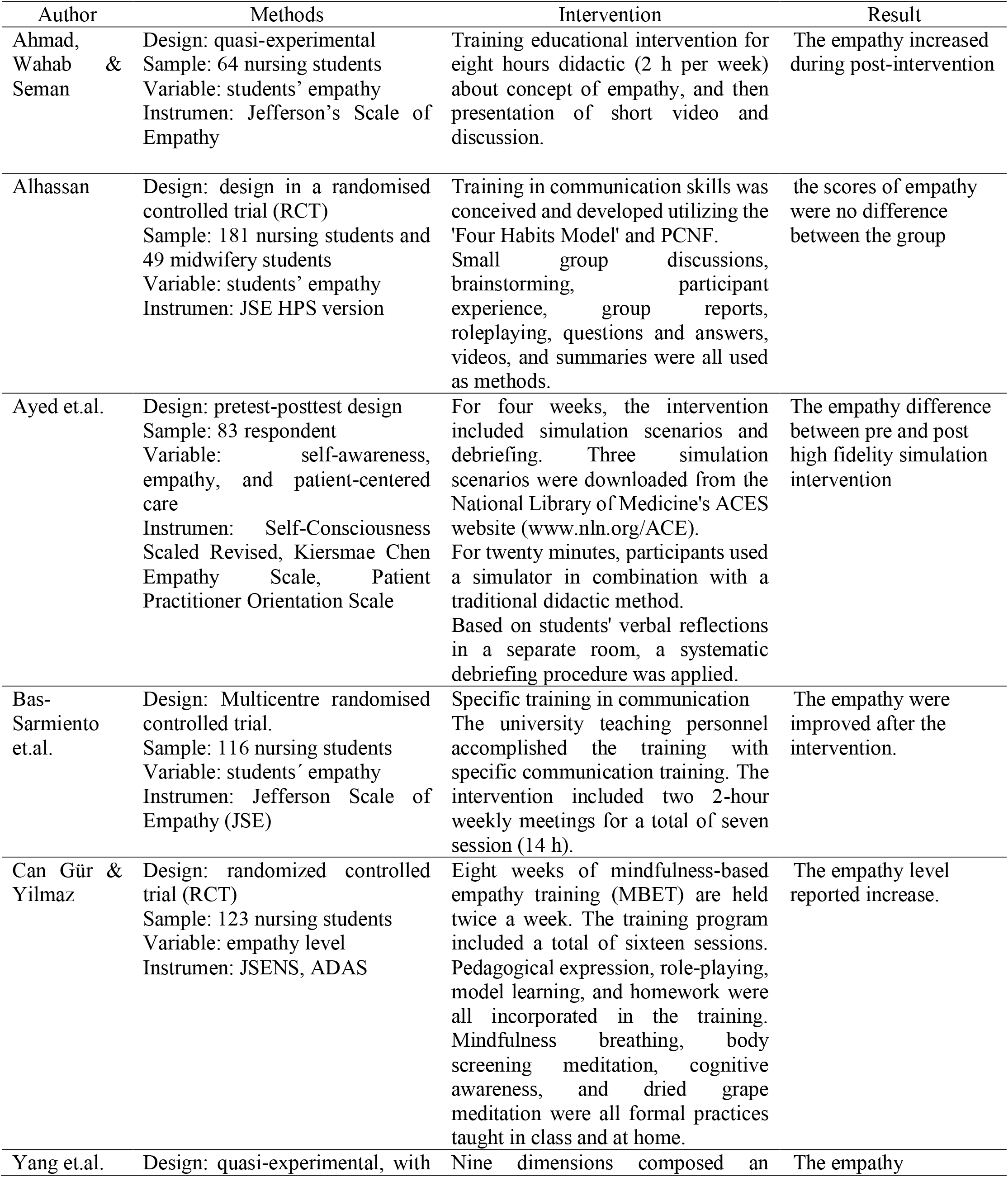
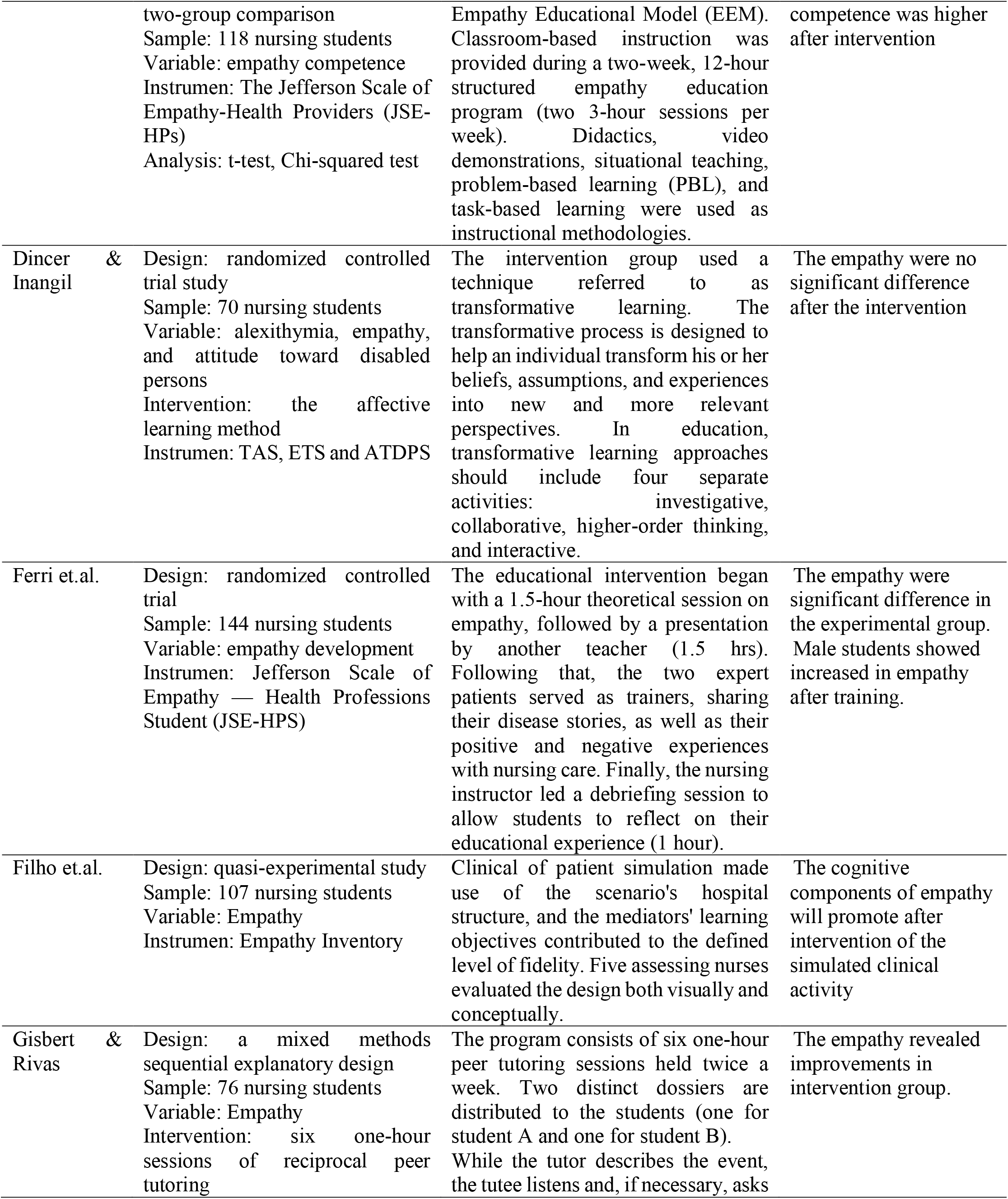
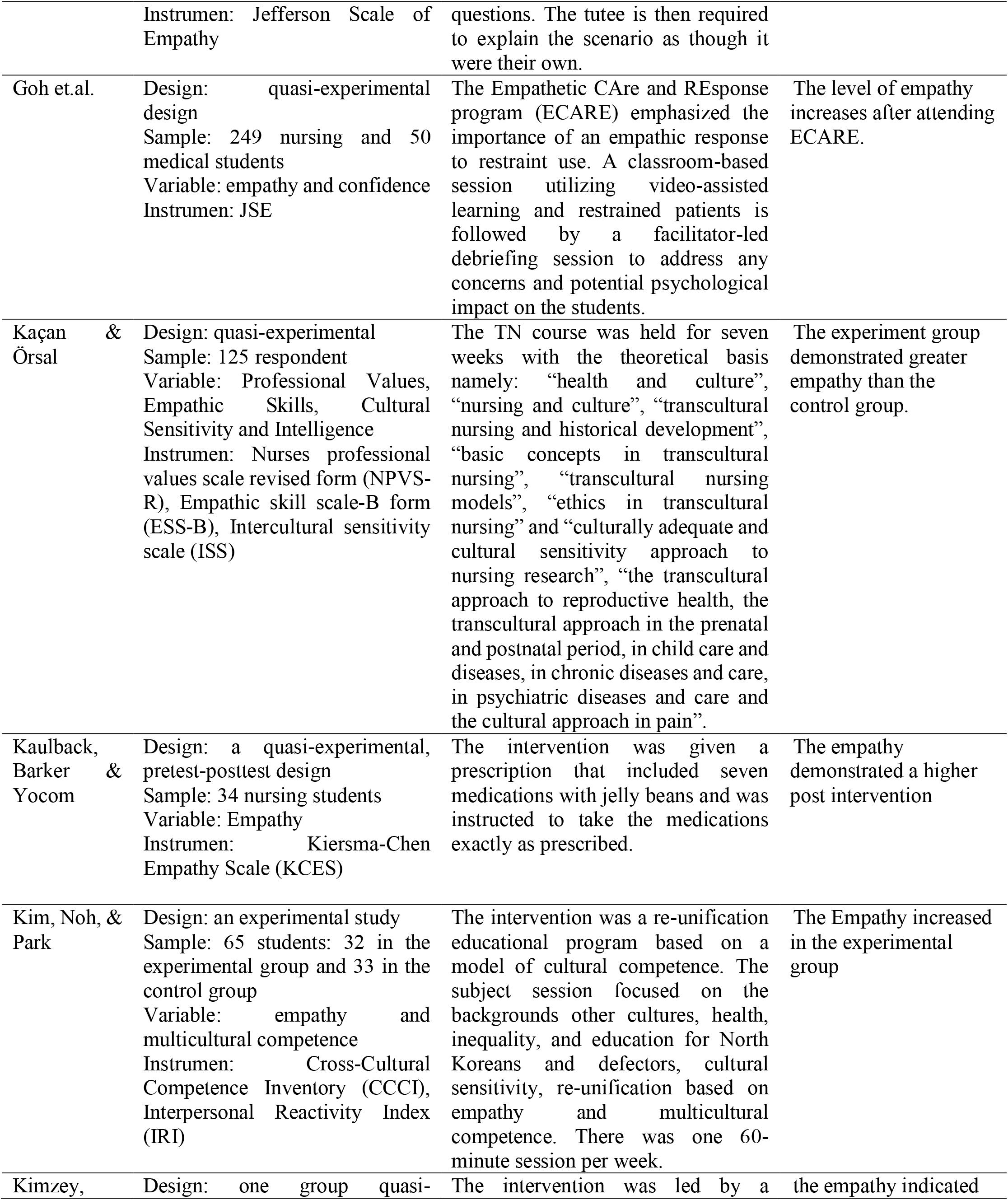
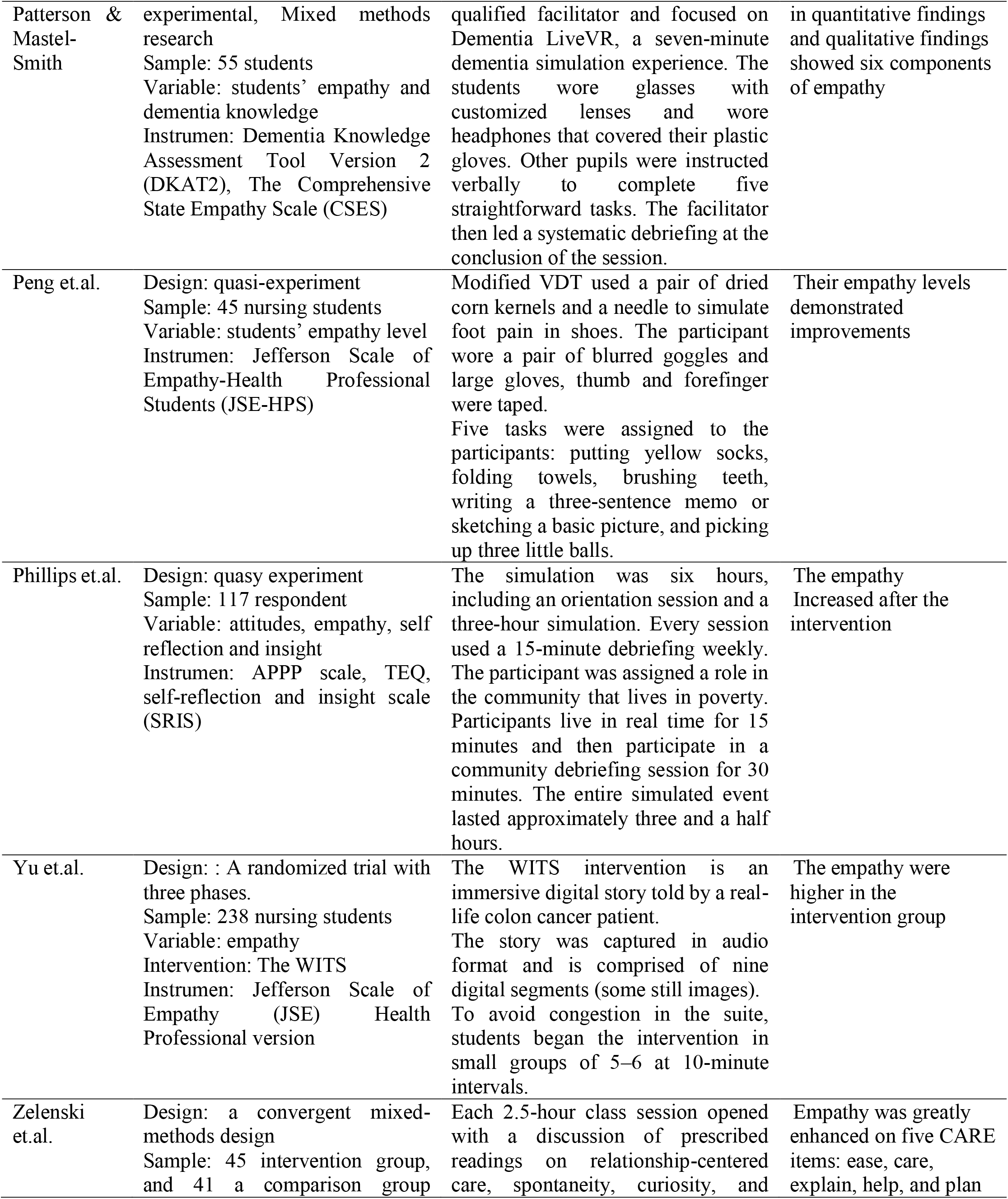
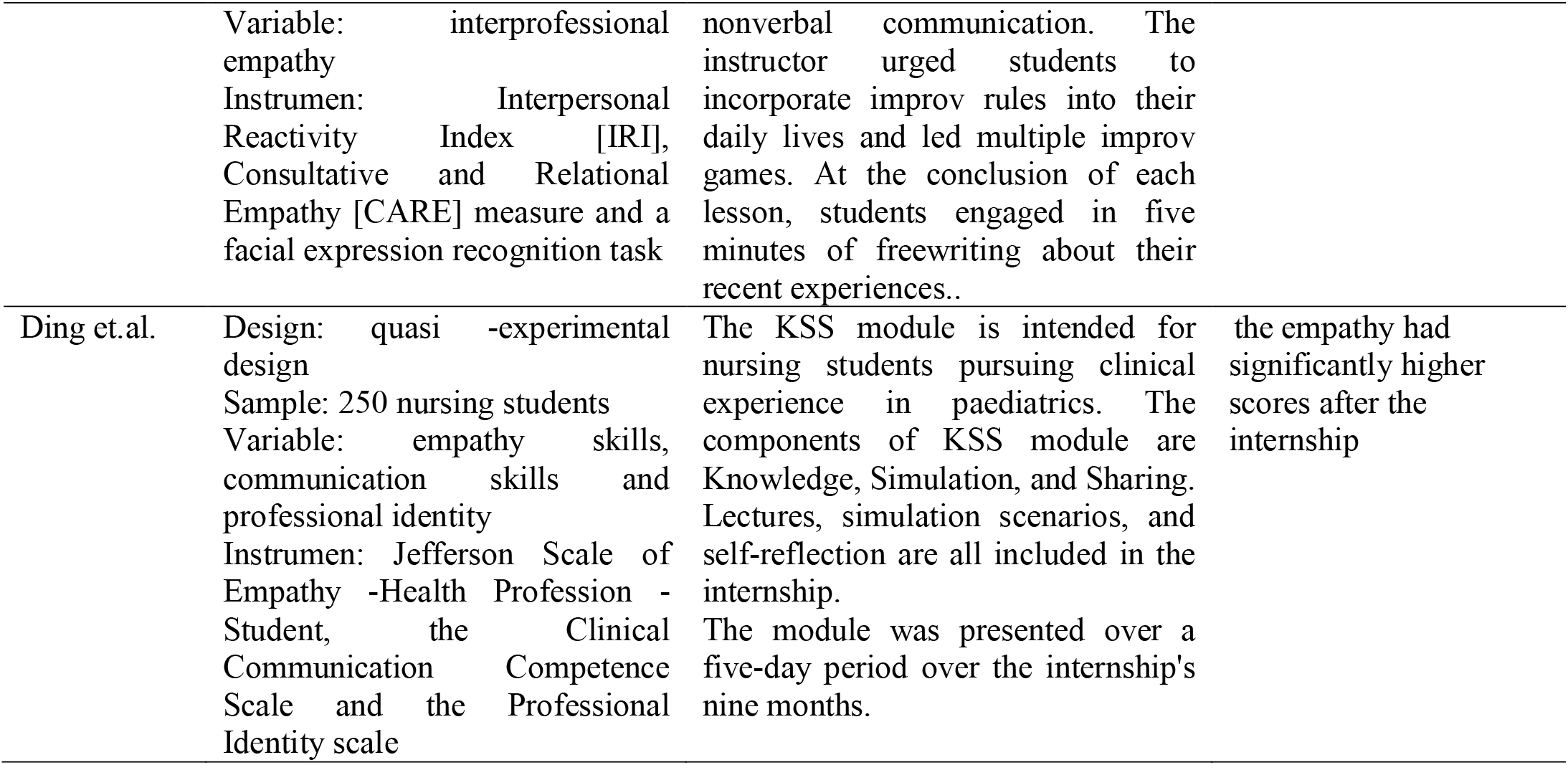
review the article

## 3. Results

### 3.1 Study Selection

Searching for articles through the database found 739 articles. After going through the selection process, 20 articles were obtained that would be reviewed (figure 1).

**Figure 1.**
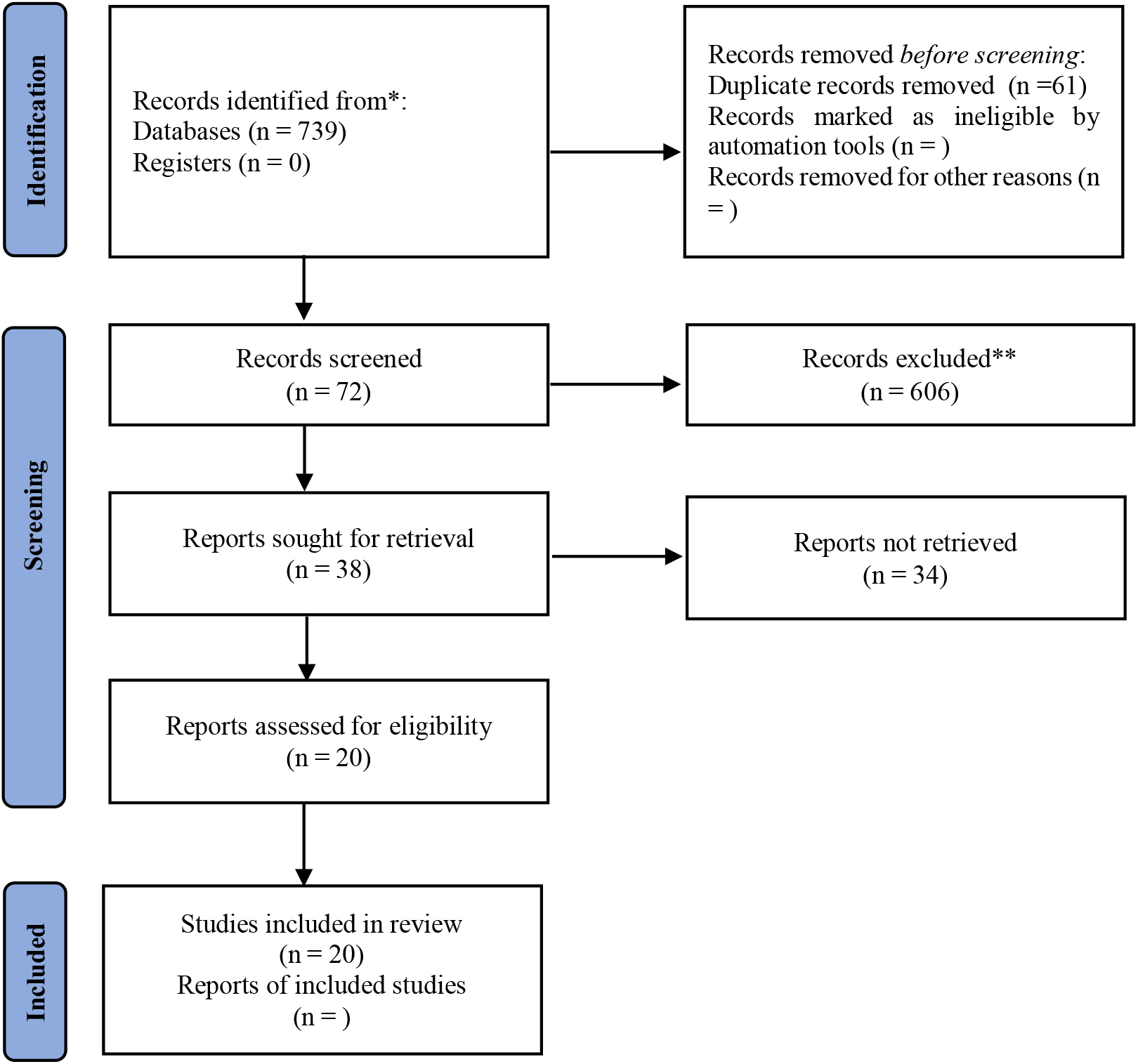
searching literature

### 3.2 Characteristics of Included Studies

The selected articles were published in 2019-2021, as many as four in 2019, 11 in 2020, and 5 in 2021. The research was carried out in 12 countries, namely four articles in the USA, three articles each in China, and Turkey, two articles in Spain, and one each from Malaysia, Ghana, Palestine, UK, Italy, Brazil, Singapore, and South Korea. The designs used in the research articles were a quasi-experiment with 13 articles, randomized control trial with six articles, and a mixed-method (with quasi-experiment) with 1 article. Based on the distribution of respondents, they were divided into one group of 5 articles and two groups of 15 articles. The measurement of respondents has measured post-test as many as two articles and pre-post test as many as 18 articles.

### 3.3 Participant Characteristics

The sampling method used convenience sampling of as many as 13 articles and random sampling as five and RCT 2 articles. The smallest sample size is 34 respondents, and the most significant sample is 250 respondents.

### 3.4 Types of Outcome Measures

The instruments to measure the empathy of nursing students used were: Jefferson’s Scale of Empathy with five articles, JSE HPS version with six articles, Kiersmae Chen Empathy Scale with two articles, Empathic Tendency Scale (ETS) with 1 article, Empathy Inventory with 1 article, Empathic skill scale-B form (ESS-B) with 1 article, Interpersonal Reactivity Index (IRI) with two articles, The Comprehensive State Empathy Scale (CSES) with 1 article, and Toronto Empathy Questionnaire (TEQ) with 1 article.

### 3.5 Empathy Outcomes

Research shows that education and training can increase student empathy in several ways.

#### Subject

Training materials that can increase student empathy are:

1. The concept of empathy through training for 8 hours where every week for 2 hours (Ahmad et al., 2020)
2. Interpersonal Communication Skills, given for 14 hours where every week for 2 hours (Bas-Sarmiento et al., 2019)
3. Mindfulness-based empathy training (MBET) teaches about Aging, communication, empathy, mindfulness, body screening meditation, acceptance and allow, awareness of the judiciary, and Effective/empathic/active listening. Training given for 16 sessions, every week carried out for two sessions (Can Gür & Yilmaz, 2020)
4. Empathy Educational Model (EEM) contains primary material of empathy, emotion and language skills, nonlinguistic skills, and practicing empathy, given for 12 hours every week 2-3 times (Yang et al., 2020)
5. Transcultural nursing education held with subject health, nursing, culture, transcultural nursing, cultural sensitivity, and nursing research, for seven weeks (Kaçan & Örsal, 2020)
6. A re-unification educational program containing a model of cultural competence, given for 1 hour 6 weeks (Kim et al., 2020)

#### Patient

Education and training using patients/patient simulations to help increase student empathy, namely:

1. Expert patients shared stories when illness, the experience of nursing care. This program was held for one day (7 hours) (P Ferri et al., 2019)
2. The Empathetic CAre and Response program (ECARE) use restrained like patients in the healthcare setting, 1-hour lecture (Goh et al., 2020)
3. Polypharmacy effects used seven medications with jelly beans, managed multidrug and multi-dose regime for seven days (Kaulback et al., 2021)
4. Clinical Simulation like Hospitalized Patient used disposable diapers in the elimination needs (Filho et al., 2020)
5. Dementia LiveVR is a simulation experience to immerse life with dementia for seven minutes (Kimzey et al., 2020)
6. Virtual Dementia Tour, participants wore three small balls, a pair of blurred goggles, and large gloves, with the thumb and forefinger of the dominant hand taped together, as well as the last three fingers of the non-dominant hand, and the VDT room was poorly light and decorated (Peng et al., 2020)
7. Games to promotes team interactions in the clinical setting for 6-8 weeks every 2.5 hours (Zelenski et al., 2020)
8. The KSS module is done for nursing students who practice in the pediatric room. The components of KSS module are Knowledge, Simulation and Sharing. The internship includes lectures, simulation scenarios and a self-reflection. The module was delivered over five days during the nine-month internship (Ding et al., 2020).

Education and training exercises using 1) mannequins through simulators’ nave, integrating simulator experience with traditional didactic approach. The intervention is done in the laboratory for four weeks (Ayed et al., 2021); 2) Peer Tutoring, after the tutor describes the incident, the tutee must describe the issue (Gisbert & Rivas, 2021); and 3) Community; Participants are assigned to a poor family and must live in their order for a simulated month separated into four consecutive weeks (Phillips et al., 2020).

## 4. Discussion

The results of this review illustrate that education and training are carried out to increase nursing students’ empathy. Of the 20 articles, 18 articles showed that education or training could increase/there was a positive relationship with the level of empathy. The education and training are about the concept of empathy, Interpersonal Communication Skills, Mindfulness-based empathy training (MBET), transcultural nursing education, and cultural competence. The method uses patients/patient simulations, peers, and the community.

Simulation methods with actual patients or like patients by humans or mannequins are proven and long-standing methods that effectively increase student empathy. The results of another review show that the most effective method is immersion and experiential Simulation in vulnerable groups and providing opportunities for reflection (T Levett-Jones et al., 2019). Simulation may be an appropriate educational methodology for developing empathy and empathy behavior (Bearman et al., 2015). A hospital simulation can provide an experience of dealing with illness and hospital life, feeling dependent and the need for contact, and dealing with being a patient. Other research shows that can increase empathetic behavior by listening to the patient’s voice as a consumer of health services (Heidke et al., 2018).

Simulation of vulnerable groups in the community can increase student empathy. Social simulations with the poverty conditions of (Menzel et al., 2014) and persons with disabilities (Tracy Levett-Jones et al., 2017) can be helpful and used in learning. Simulations with mannequins can increase student empathy following the thesis and goals that NLN has made (Haley et al., 2017).

Educational materials and lectures given to students to increase empathy are also reviewed, namely communication skills, interprofessional skills, and the concept of empathy (Batt-Rawden et al., 2013), and cultural and linguistic differences through three-dimensional simulations (Everson et al., 2015).

Based on the material provided, the length of education or training carried out varied from 6 hours (Kim et al., 2020), 8 hours (Ahmad et al., 2020), 12 hours (Yang et al., 2020), and 14 hours (Bas-Sarmiento et al., 2019). Education and training time is carried out starting from 1 day (P Ferri et al., 2019), four weeks (Ayed et al., 2021), six weeks (Kim et al., 2020), seven weeks (Kaçan & Örsal, 2020), and eight weeks (Can Gür & Yilmaz, 2020; Zelenski et al., 2020). On average, each week face-to-face for 2 hours. The time determination is adjusted to the material discussed and according to the stages of the learning. It guides lecturers and faculties in planning education and training for nursing students.

### Implications for education

The experiential learning method with patient simulation becomes like a patient, mannequin, and immersion in the community and helps increase student empathy. The experiential learning method with Simulation and immersion can be used as a learning method in nursing lectures or training.

## 5. Conclusion

Nursing students’ empathy can be increased by improving cognitive, affective, and psychomotor aspects. Education and training can improve cognitive aspects by providing knowledge about empathy, communication, meditation, and cultural competence. Simulation and immersion with patients, being like patients, using mannequins, or interacting with vulnerable groups improve students’ affective and psychomotor aspects.

## Data Availability

All data produced in the present work are contained in the manuscript

